# Deoxycholic acid liganded HBs contributes to HBV maturation

**DOI:** 10.1101/2021.12.21.21268182

**Authors:** Yuxue Gao, Qiqi Ning, Pengxiang Yang, Yulin Zhang, Yuanyue Guan, Ning Liu, Haijing Ben, Yang Wang, Mengcheng Liu, Tongwang Yang, Yuying Cai, Zhongjie Hu, Mengxi Jiang, Dexi Chen

**Author notes:** These authors contributed equally to this work (Yuxue Gao, Qiqi Ning, Pengxiang Yang, Yulin Zhang).

## Abstract

Understanding the underlying mechanism of HBV maturation and subviral particle production is critical to control HBV infection and develop new antiviral strategies. Here, we demonstrate that deoxycholic acid (DCA) plays a central role in HBV production. HBV infection increased DCA levels, whereas elimination of DCA-producing microbiome decreased HBV viral load. DCA can bind to HBs antigen via LXXLL motif at TM1 and TM2 region to regulate HBs-HBc interaction and the production of mature HBV. Plasma DCA levels from patients undergoing antiviral therapy were significantly higher in those with positive HBV viral load. These results suggest that intestinal DCA-producing microbiome can affect the efficiency of antiviral therapy and provide a potential novel strategy for HBV antiviral therapy.

**One Sentence Summary:** We demonstrate that DCA-promoted HBs-HBc interaction and contributes to HBV maturation.

---

Hepatitis B virus (HBV) surface proteins (HBs) are initially synthesized as multispanning membrane proteins of the endoplasmic reticulum (ER) membrane and constructed a defined transmembrane (TM) topology (*1, 2*). Small hepatitis B virus surface antigens (SHBs) are inserted into the ER membrane via various TM regions. The first ER TM region (M1) encompasses aa 8-22 and the second TM region (M2) encompasses aa 80-98 (*3-5*). HBV particle enveloping requires HBs-HBc interaction (*6*). Interestingly, there is an LXXLL motif in the ER side of the HBs M1 and 3 LXXLL motifs in the HBs M2, which is also a typical binding site for bile acids (*7*).

Here, we first identified high levels of DCA in HepG2.2.15 cells, which contain integrated HBV DNA genome and produce HBV pgRNA and mature viral particles (*8*), as compared with HBV-free HepG2 cells. HBV-infected HepG2-NTCP cells that express human sodium taurocholate co-transporting polypeptide (NTCP) also contain higher DCA content compared with non-infected HepG2-NTCP cells (Fig. 1A) (*9*). DCA treatment increased HBV viral load released from HepG2.2.15 cells (Fig. 1B) and HBs level in HepG2-NTCP cells (Fig. 1C). We further compared the hepatic secondary bile acid levels of the HBV transgenic mice (HBV Tg) and control mice (C57BL/6), and found higher DCA levels and lower LCA levels in the livers of HBV Tg mice (Fig. 1D). In addition, the hepatic DCA levels were positively correlated with the HBV DNA copies of the plasma in the HBV Tg mice (Fig. 1E). Then, we placed the HBV Tg mice on antiviral therapy with Entecavir (ETV) at 0.1 mg/kg/day and found a significant decrease of HBV DNA copies in the plasmas and liver tissues of the mice one week after treatment (Fig. 2A, 2B). Interestingly, hepatic DCA level of the HBV Tg mice was also parallelly decreased one week after ETV treatment (Fig. 2C). In contrast, the hepatic LCA level was increased following ETV treatment (fig. S1). These results indicate that plasma HBV viral load was positively correlated with hepatic DCA levels. Similar results were identified from *in vitro* studies with the HepG2.2.15 cells (fig. S2).

**Fig. 1.**
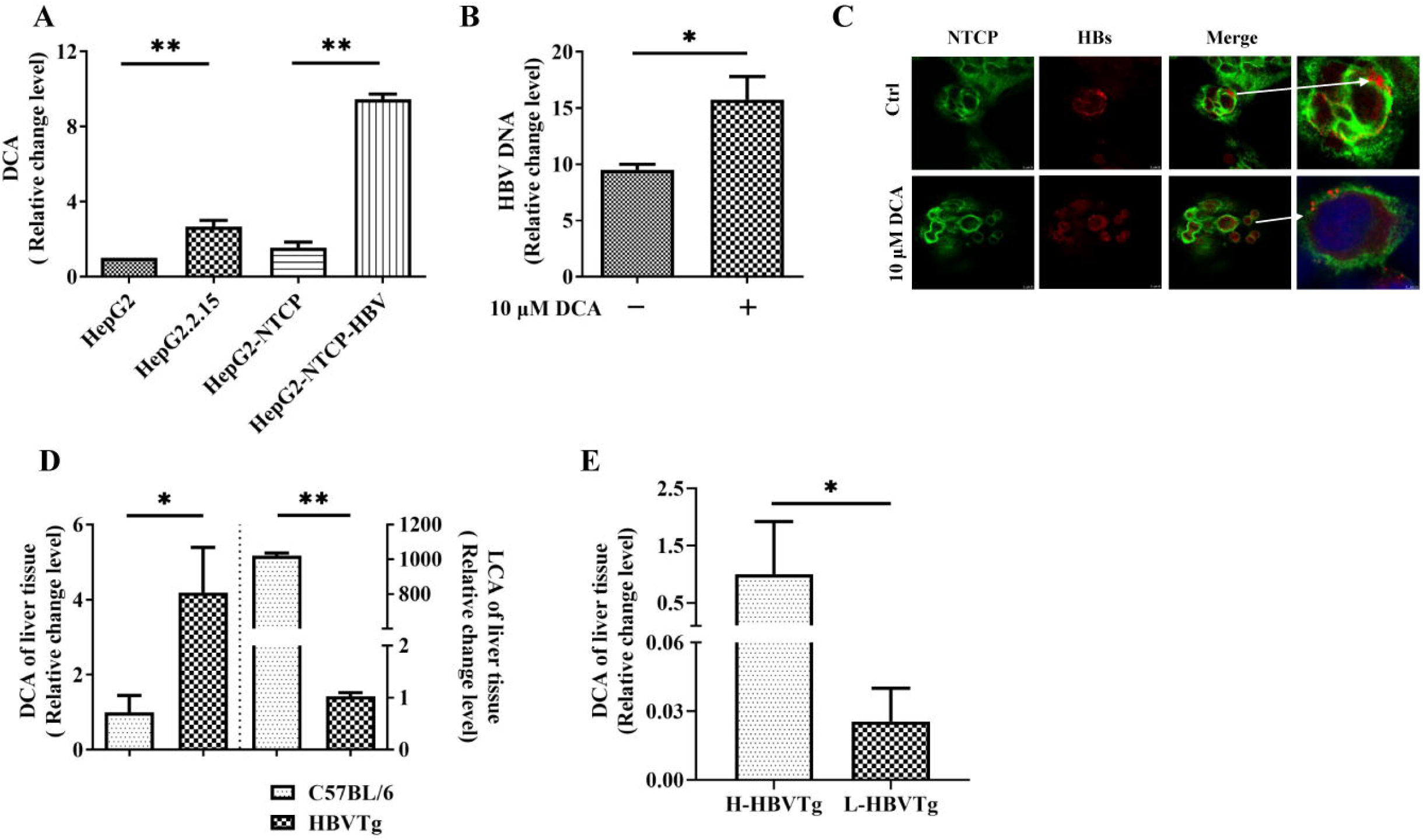
The effect of DCA on HBV viral load *in vivo* and *in vitro*. (A) Relative fold change level of DCA in HepG2, HepG2.2.15, and HepG2-NTCP cells with or without HBV infection (n=3). (B) Relative fold change level of HBV DNA in HepG2.2.15 cells treated with 10 μM DCA (n=3). (C) IF of HBsAg in HBV-infected HepG2-NTCP cells treated with 10μM DCA for 5 days. Scale bar, 10 μm. (D) Relative fold change levels of hepatic DCA and LCA in HBV Tg mice and C57BL/6 mice (n=12). (E) Relative fold change level of hepatic DCA in HBV Tg mice with high (H-HBV Tg) or low (L-HBV Tg) plasma HBV DNA copies (n=5). \**P*<0.05, \*\**P*<0.01.

**Fig. 2.**
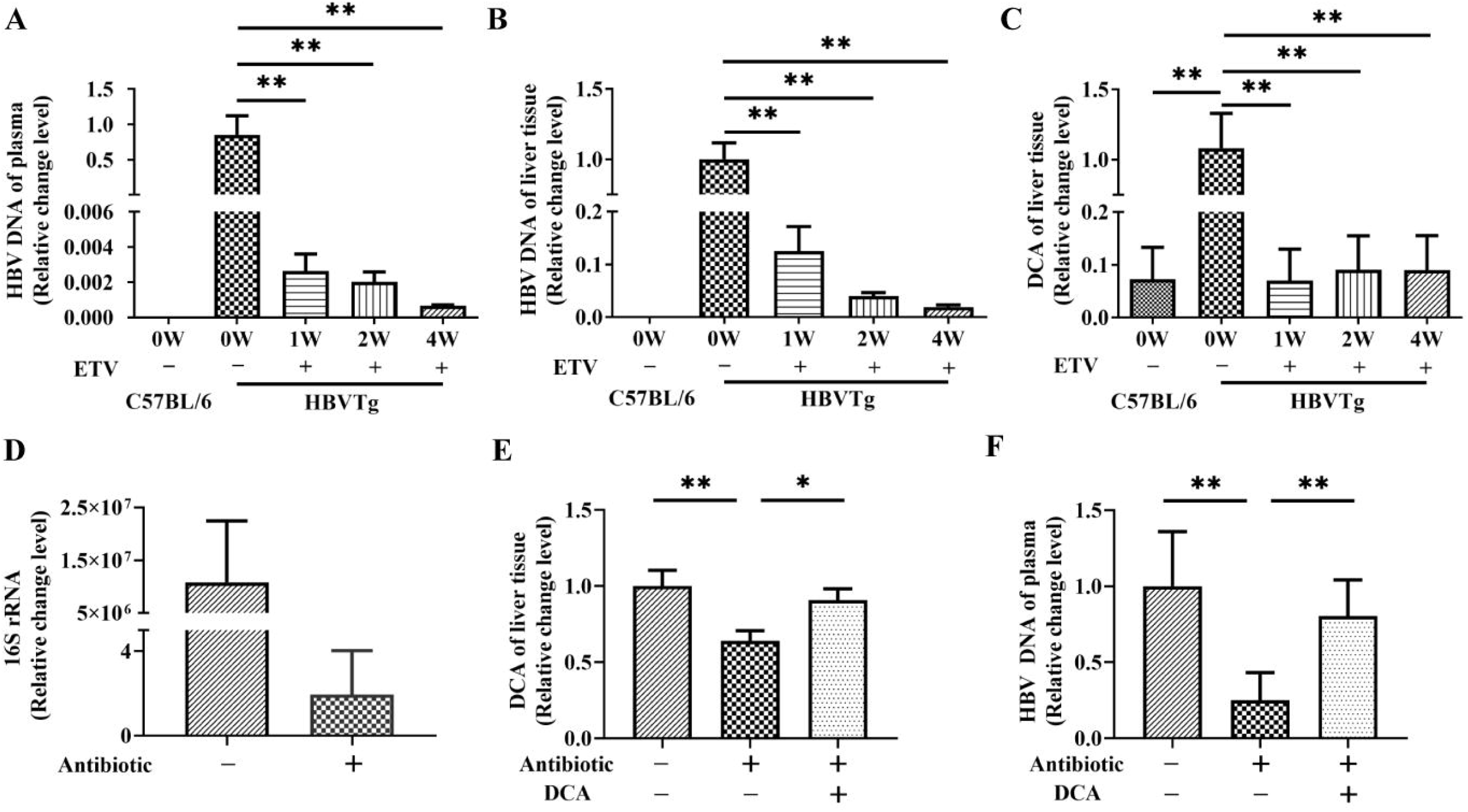
HBV viral load is closely associated with DCA level in HBV Tg mice treated with ETV and antibiotics. (A) and (B) Relative fold change levels of plasma HBV DNA (A), hepatic HBV DNA (B), and hepatic DCA (C) in HBV Tg mice treated with ETV for 1 to 4 weeks (n=7). (D) Relative fold change level of gut microbiota-derived 16S RNA in antibiotics-treated HBV Tg mice was calculated using the 2^−ΔΔCT^ method (n=4). (E) Relative fold change level of hepatic DCA in HBV Tg mice treated with antibiotic and DCA (n=4). (F) Relative fold change level of plasma HBV DNA in HBV Tg mice treated with antibiotic and DCA (n=4). \**P*<0.05, \*\**P*<0.01.

After observing the association between HBV viral load and hepatic DCA levels, we next fed HBV Tg mice with antibiotics polymyxin B (25-35mg/kg/day) and vancomycin (125-175mg/kg/day) for fourteen days to eliminate intestinal microbiomes, from which DCA is produced. Fecal 16S rRNA levels were analyzed to monitor the efficiency of intestinal sterilization (Fig. 2D). The hepatic DCA levels and the plasma HBV DNA levels were decreased in antibiotic-treated HBV Tg mice and recovered following the DCA feeding HBV Tg (Fig. 2E, 2F). These results indicate that the hepatic DCA level of HBV Tg mice strongly affects their plasma HBV viral load.

The envelopment of HBV requires the interaction of both the preS1 domain (PLSPPLRNTHPQAMQWNSTTF) and S domain (PTSNHSPTSCPPTCPGYRNMCCRRF) with HBc (*10, 11*). However, the detailed mechanisms of HBV-HBc interaction are still unclear. There is an LXXLL (LGPLL) domain in HBs M1 region and 3 LXXLL (LFILLLCLIFLLVLL) domains in the HBs M2 region. As amphiphilic molecules, bile acids may interact with HBs’s TM domain to affect HBs’ structure. To identify the role of HBs LXXLL domains in the formation of intact HBV particles, we constructed M1 and M2 mutated plasmids (Fig. 3A) and measured the viral load from the culture supernatant of transfected HepG2 cells. Compared with HBV-WT plasmid-transfected HepG2 cells, the culture supernatant viral load was higher in HBV-M1 group and lower in HBV-M2 group (Fig. 3B). The infection and production capacity of the mutated virus were detected in HepG2-NTCP cells by immunofluorescence targeting HBs three days post-infection (Fig. 3C). The results showed that the HBs antigen in HBV-WT infected cells was highly expressed and presented in particle form, which were lost in HBV-M1 or HBV-M2 infected HepG2-NTCP cells. The *in vitro* results were confirmed by plasma HBV DNA, hepatic HBs and DCA from wildtype and M1, M2 mutated HBV Tg mice (table S1). The results showed that the infection viability and viral load of M1 and M2 HBV were significantly lower than those of HBV-WT, indicating the role of M1 and M2 domain in HBV maturation.

**Fig. 3.**
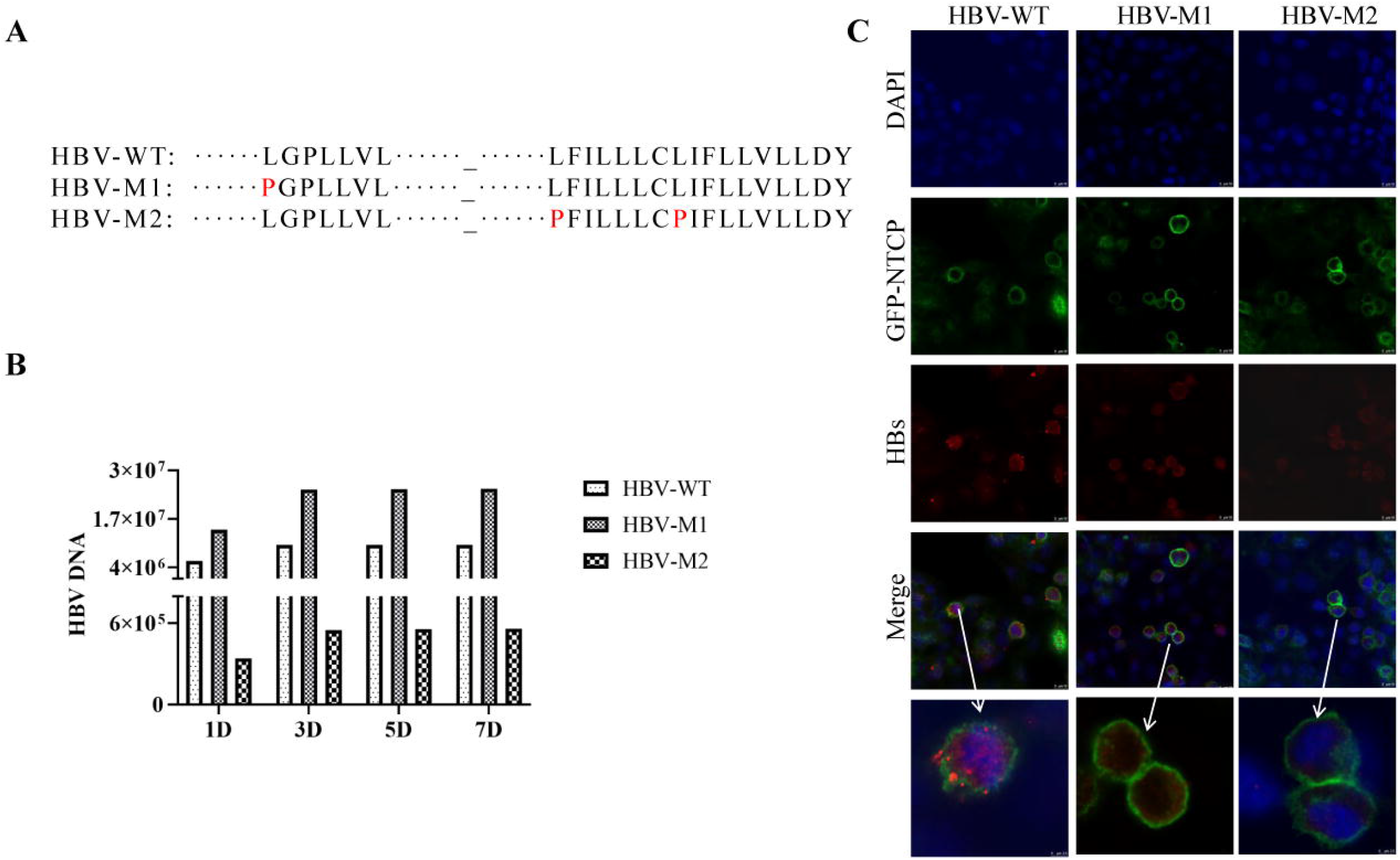
Bile acid binding motif (LXXLL) in TM1 (LGPLL) and TM2(LFILLLCLIFLLVLL) of S domain determines HBV release and intracellular transport. (A) The M1- and M2-mutation sites in HBV expression vector. (B) The HBV viral loads in culture supernatant at different time points following transient transfection of wildtype and mutant HBV expression vectors in HepG2 cells. (C) IF of HBsAg in HBV-infected HepG2-NTCP cells following equal amount of wild-type (WT), M1, and M2 culture supernatant HBV DNA infection. Scale bar, 10 μm.

To verify whether DCA participates in HBs-HBc interaction, we ran the Immunoprecipitation (IP) using HBV-WT, HBV-M1, or HBV-M2 mice. The immune-blot results showed that HBs and HBc could be detected by WB when 5Ð10^7^ copies of viruses were loaded (Fig. 4A). By performing IP with anti-HBc antibody, we found that the HBc-HBs interaction was significantly decreased in HBV-WT mice following antibiotics treatment and recovered by DCA feeding (Fig. 4B). Then we explored the role of LXXLL motif in DCA binding and HBc-HBs interaction. ELISA assay showed a significantly higher HBs expression in HBV-M2 transgenic mice than in HBV-WT and HBV-M1 transgenic mice (Fig. 4C). However, the hepatic DCA level in HBV-M1 mice is significantly higher than that in HBV-WT mice and HBV-M2 mice (table S1). The IP was performed by adjusting the weight of liver tissues with HBs level matching 10^8^ copies of HBV DNA (Fig. 4D). The results showed that compared with HBV-WT and HBV-M2 mice, the binding capacity of hepatic HBc to HBs is significantly higher in HBV-M1 mice, which also had higher plasma HBV viral load and hepatic DCA levels (table S1). In contrast, even with a high level of hepatic HBs, HBV-M2 mice, which had a low-level hepatic DCA, showed a low plasma viral load and a low level of HBs-HBc interaction (Fig. 3B, 4D). The above results suggest that hepatic DCA contributed to the HBs-HBc interaction *in vivo*. The LXXLL motif in TM1 and TM2 of HBs is a typical binding site of bile acids (*12*). TM1 of the S domain locates in the cytosolic compartment, while TM2 anchors in the lipid bilayer of HBs (*11*). Unlike TM1, TM2 mutation that lost the ability to interact with DCA may change its anchoring distance in the lipid bilayer of HBs (fig. S3), which likely changes HBs-HBc binding ability.

**Fig. 4.**
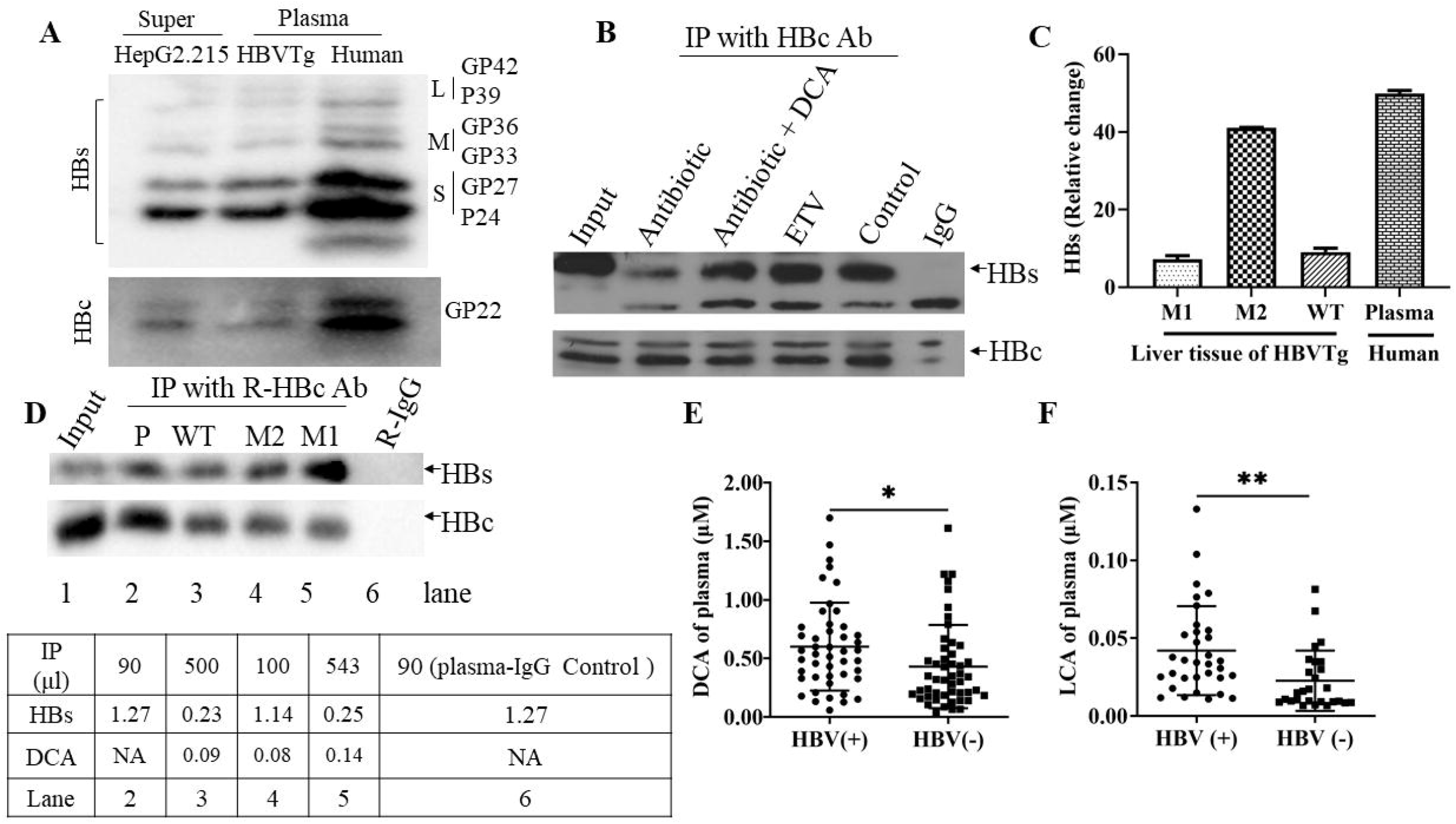
DCA-regulated HBc-HBs interaction in HBV Tg mice and HepG2.2.15 cells; DCA and LCA level in the LLV and LDL patients following 54 weeks of NA treatment. (A) HBs and HBc were detected by WB to confirm HBV loads. (B) Involvement of DCA in regulating HBs-HBc interaction was detected following antibiotic, ETV and DCA treatment. (C) Relative fold change level of hepatic HBs in different mutated and wild-type HBV Tg mice as determined by ELISA. (D) The HBc-HBs interaction is associated with DCA level in liver tissue of HBV Tg mice. Plasma levels of DCA (E) and LCA (F) in the patients with or without LLV following 54 weeks of NA treatment. *\*P*<0.05, *\*\*P*<0.01.

The maintenance of low-level viremia (LLV) in some chronic HBV-infected patients continually causes chronic liver damage and challenges antiviral therapy (*13-15*). Serum bile acids have been described as prognostic markers to predict anti-HBV treatment outcomes in chronic HBV-infected patients (*16*). A total of 111 chronic HBV-infected patients who maintained over 54 weeks of antiviral therapy with nucleoside analogs were selected for this study at Beijing Youan Hospital. Among them, 53 LLV patients’ HBV load remained about 1000 copies per milliliter, and the other 58 patients’ viral load was below the lower detection limit (LDL). The detailed clinical data are shown in table S2. Bile acid levels in these patients were analyzed by mass spectrometry. The results showed that the patients with LLV had higher plasma DCA and LCA concentrations, but not other bile acid concentrations, than those with HBV load below LDL (Fig. 4E and 4F, fig. S4). Considering the high level of hepatic DCA in HBV Tg mice, we speculate that the increased plasma DCA in chronic HBV-infected patients with LLV may be related to the increase of secondary bile acid produced by intestinal flora. These clinical results further support the hypothesis that DCA was involved in HBV production. Results from antibiotic treatment also provide a potential direction for HBV antiviral treatment.

## Data Availability

All data produced in the present study are available upon reasonable request to the authors

## Acknowledgments

We thank Y. Huang, X. Liu, X. Guo and L. Qiao for laboratory assistance.

## Author contributions

Conception and design: DXC, MXJ, YLZ, ZJH

Methodology: YXG, QQN, PXY, YYG, NL, HJB, YW, YYC

Visualization: YXG, QQN, PXY, YYG

Writing-original draft: YXG, DXC, MXJ, YLZ, MCL, TWY

Writing-review & editing: YLZ, MXJ, MCL, DXC

## Data and materials availability

All data are available in the main text or the supplementary materials.

## Supplementary Materials

Materials and Methods

Figs. S1 to S4

Tables S1 to S2

