## Supplementary Materials for "Deoxycholic acid liganded HBs contributes to HBV maturation"

This file includes:

Figs. S1 to S4

Tables S1 to S2

**Materials and Methods**

**Cell culture, molecular cloning and transfection of HBV constructs, HBV infection and drug treatments**

HepG2 and Hep2.2.15 cells were maintained in modified eagle medium (MEM) (Gibco, cat# 61100103). HepG2-NTCP cells were maintained in MEM supplemented with 0.5 mg/mL G418 (Invitrogen, cat#11811031). All cell media were supplemented with 10% fetal bovine serum (FBS) (Gibco, cat# 10270-106), penicillin (100 U/mL), streptomycin (100 µg/mL) (Gibco, cat# 15140122). Cells were maintained at 37ºC with 5% CO2.

A replication-competent HBV clone (WT) was constructed by plasmids containing 1.1 copies of the HBV genome under the control of a CMV promoter. In this study, we constructed two HBV mutant plasmids, namely M1(T1575C) and M2 (T1800C, T1821C). Briefly, the plasmids for WT, M1, and M2 were transfected into HepG2 cells with X-tremeGENE HP DNA Transfection Reagent (Roche, cat# 6366236001). WT-, M1-, and M2-HBV secreted in culture medium were harvested every day after transfection.

Cell culture-generated HBV was obtained by the transfection of different HBV constructs. The culture supernatant was centrifuged at 11000g for 30 min to remove cell debris, concentrated with Amicon Ultra centrifugal filters (100 kDa, Merck Millipore, cat# UFC910096). The purified cell culture-generated HBV was inoculated onto HepG2-NTCP cells.

10 µM of DCA (Sigma-Aldrich, cat# D2510) and Entecavir were used in this study unless otherwise indicated.

**Immunofluorescence (IF) assay**

For IF, cells were washed twice with sterile 1×PBS and fixed with 4% paraformaldehyde for 10 minutes. Samples were washed with phosphate-buffered saline (PBS) three times, permeabilized with 1% TritonX-100 for 20 minutes, then washed again with sterile 1× PBS three times. Cells were then incubated in blocking buffer (1% goat serum in 3% bovine serum albumin) for 1 h at 37ºC. Cells were incubated with mouse anti-HBcAg antibody or mouse anti-HBsAg antibody (kindly provided by Xiamen University, China) diluted in 3% bovine serum albumin in sterile 1×PBS at 4ºC overnight, repeatedly washed, and subsequently incubated with anti-mouse-IgG conjugated with Alexa Fluor 594 (Jackson ImmunoResearch, cat# 109-585-043). After performing nuclear staining was with 4',6-diamidine-2-phenylindole (DAPI, Sigma Aldrich, cat# D9542), cells were examined with confocal microscopy.

**Detection of HBV viral load**

HBV DNA was extracted from fresh liver tissue, plasma, and cell culture supernatant. HBV DNA viral loads were determined using the Abbott RealTime HBV viral load assay with 0.5ml DNA extraction protocol (Abbott Molecular, USA).

**Immunoprecipitation assay and Western blot analysis**

Plasma and cell culture supernatant were centrifuged at 110, 000 g for 70 minutes. HBV ultracentrifugation products or liver tissues were resuspended in 500 μl lysis buffer (20 mM Tris, pH 7.5, 150 mM NaCl, 1.0% Triton X-100, 0.5% Nonidet P-40, 1 mM EDTA and protease inhibitor). Immunoprecipitation assays were performed by incubating 1000 μg of protein lysates with Hepatitis B Virus Core Antigen (Abcam, cat# ab115992) at 4℃ overnight. Next, the samples were incubated with protein A/G beads (Topbio, cat# P1040) at room temperature for 2 hours, according to the manufacturer’s instructions. After beads were washed 4 times with the wash buffer, samples were eluted with 2*SDS-PAGE loading buffer (with β-Mercaptoethanol) and denatured by incubation at 100℃ for 5 min. Protein lysates (10% of the starting material used for immunoprecipitation) and immunoprecipitates were separated by 12% SDS–PAGE gels and transferred to nitrocellulose membranes. Membranes were incubated with indicated primary antibodies, washed, and probed with HRP-conjugated secondary antibodies before subjecting to chemiluminescence (Thermo Scientific). The resulting bands were digitalized and quantified using the NIH Image J software.

The following antibodies were used in this study: anti-Hep B sAg (1:000, Santa-Cruz Biotechnologies, cat# sc-53299), anti-HBcAg (1:1000, Invitrogen cat# MA1-7609), anti-mouse secondary antibodies (1:2000, Cell Signaling Technology, cat# 7076S).

**UPLC-MS/MS profiling of bile acids**

Methanol aqueous solution was diluted into a series of standard working solutions, and the standard curves were established by the internal standard method. 50μL of supernatant from each sample with 150μl precipitator containing isotope internal standard was vortexed for 10 min, mixed with chlorine sodium solution, followed by centrifugation at 14,000 rcf for 10 min at 4 °C. The Triple Quad™ 4500MD (Sciex) mass spectrometer coupled to LC-MS/MS system was used to detect bile acids. Mobile phase A contained 5 mM ammonium acetate in water, and mobile phase B was methyl alcohol. The column temperature was set at 45 °C. A QC sample was prepared at a flow rate of 800 μL/min by aliquoting 20μL of each sample and the same QC strategy was used as above. Mass spectrometry was performed using electrospray in negative ESI ionization modes and MRM. The conditions of the mass spectrometer were as follows: source temperature was 550 °C, ion Source Gas1Gas1 55psi, Ion Source Gas2 55psi, Curtain gas 40psi; ionSapary Voltage Floating-4500 V.

**HBsAg and HBcAg ELISA**

75 μL sample was tested for HBsAg using ELISA kits from KHB (Kehua Bio-engineering Co, China) according to the manufacturer's instruction. S/CO values of ≥1.00 for HBsAg were considered reactive according to the manufacturer's instruction.

Microtiter wells (Costar, cat# 42592) were coated with 100μl of anti-HBcAg antibody (Santa Cruz, cat#sc-23947) (about 1μg/mL) with coating buffer (pH 9.6) at 4℃ overnight, and the wells were washed four times with a washing buffer (0.1% Tween-20 in PBS, pH 7.2), followed by incubation with 300μl 5% BSA blocking reagent at 37℃ for 2 h. 75μL sample dilution and the negative, positive sample was added to each well. The wells were incubated for 1 h at 37℃, then wells were washed five times with a washing buffer, followed by incubation with 100μL of HRP-conjugated anti-HBcAg solution (1:1000, Santa Cruz, cat#sc-23947 HRP) for 30 min at 37℃. Then wells were washed four times with a washing buffer and allowed to react with 100μL of substrate solution for 15 min at 37℃. The reaction was terminated by adding 50μL of 2M sulfuric acids, and the absorbance was determined at 450 nm with a microplate reader.

**Animal studies**

Hepatitis B virus transgenic mice (C57BL/6, strain HBV1.3.32) of age 4-6 weeks were treated with Entecavir at 0.1 mg/kg/day dispensed in the drinking water. Cages of mice were treated ad-lib at 4 to 6 weeks of age with 25mg/kg-35mg/kg/d Polymyxin B (MCE, cat# HY-A0248) and 125mg/kg-175mg/kg/d vancomycin (MCE, cat# HY-B0671). Polymyxin B and vancomycin were added in drinking water and were replaced every day. Mice were examined daily for health conditions and sacrificed at indicated time points, when serum as well as liver tissues were obtained. Cages, food, and water were autoclaved, and mice were handled in sterile hoods.

Animal experiments were carried out in accordance with the recommendations in the Guide for the Care and Use of Laboratory Animals of the National Institutes of Health after approval by the Institutional Animal Care and Use Committee at the Capital Medical University (Assurance number AEEI-2021-021). Mice were housed and treated according to the guidelines established by the National Institutes of Health Guide for the Care and Use of Laboratory Animals and housed in a temperature-, humidity-, and light-controlled room.

**Statistical Analyses**

Univariate analyses were performed using GraphPad Prism 9. Mann-Whitney *U* test, Student's *t*-test, Fisher’s exact test were used for unmatched comparisons. *P* values less than 0.05 were considered significant.

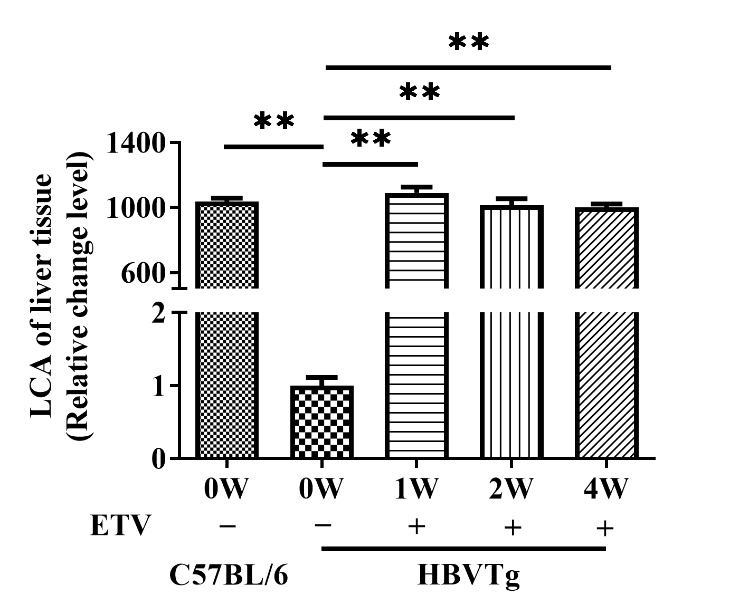

**Fig. S1. Relative fold change level of hepatic LCA in HBV transgenic mice.** Relative fold change level of hepatic LCA in HBV transgenic mice treated with ETV for 1 to 4 weeks (n=7). **P*<0.05, ***P*<0.01.

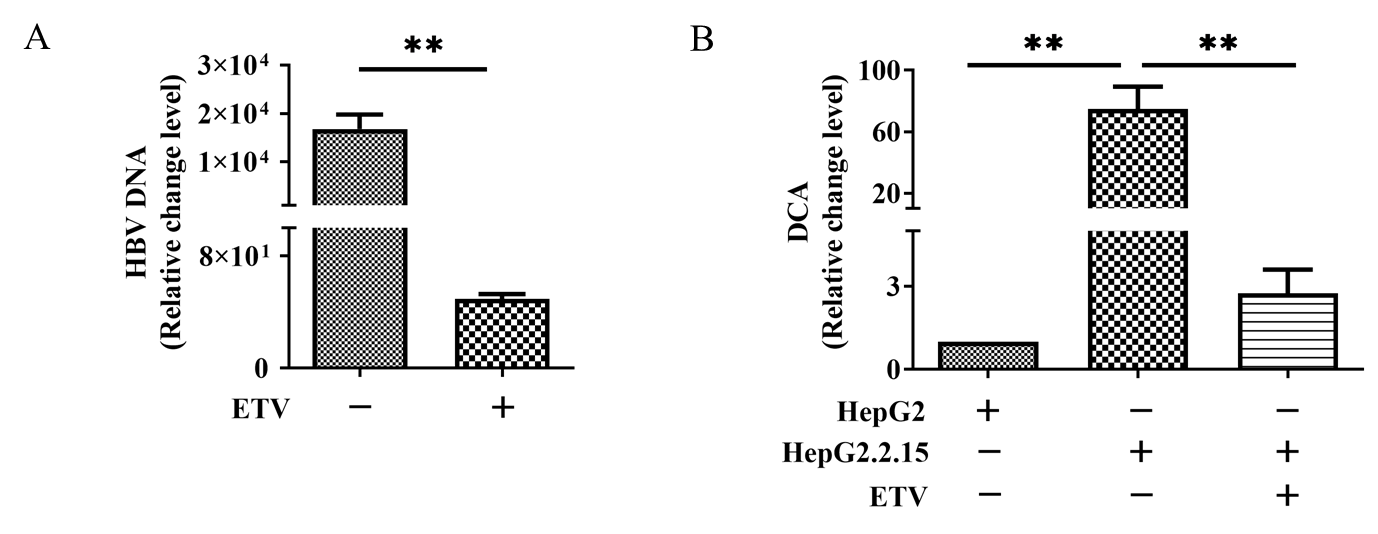

**Fig. S2. Relative fold change level of HBV DNA and DCA.** (A) Relative fold change level of HBV DNA in culture supernatant of HepG2.2.15 cells treated with ETV for 5 days (n=3). (B) Relative fold change level of DCA in HepG2 and HepG2.2.15 cells treated with ETV for 5 days (n=3). **P*<0.05, ***P*<0.01.

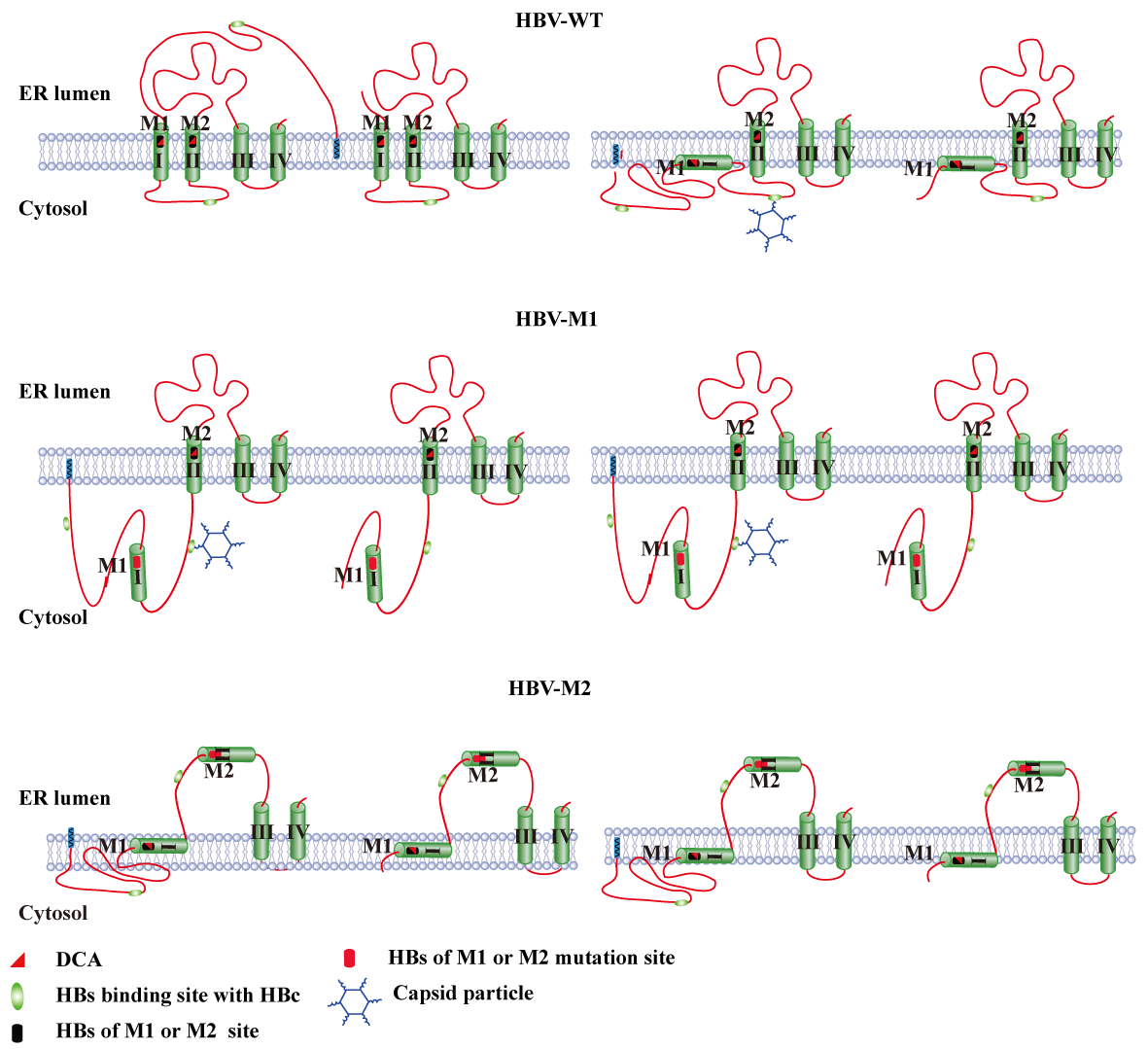

**Fig. S3. Topology of the HBV-WT envelope protein and M1- and M2-mutated proteins.** TM1 and TM2 of large HBs of S domain anchors in the HBV lipid bilayer. Unlike TM1, TM2 mutation that lost the ability to interact with DCA, may change the anchoring distance of HBs in HBV lipid membrane.

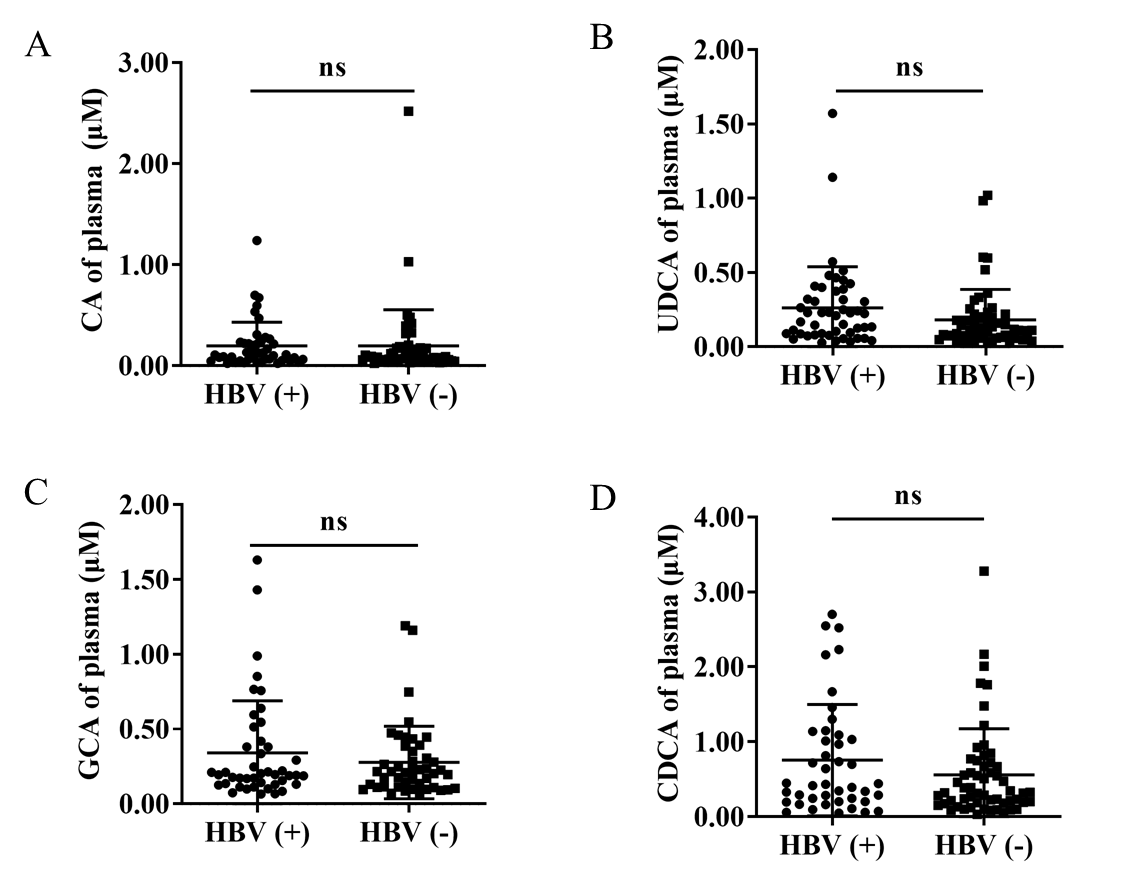

**Fig. S4. Plasma levels of bile acids in patients with LLV and LDL following 54 weeks of NA treatment.** (A)CA, (B)UDCA, (C)GCA, and (D)CDCA level in patients with LLV and LDL following 54 weeks of NA treatment.

**Table S1. Viral load of different S domains-mutated HBV in the plasma or liver tissue of mice (n=4).**

| Types | Viral loads (plasma, mean) | HBs  (liver, mean) | DCA  (liver, mean) | plasma HBV/DCA（mean） | Viral load （HepG2-NTCP, mean） |
| --- | --- | --- | --- | --- | --- |
| HBV-M2 | 2.00E+6 | 1.14 | 0.084 | 2.08E+7 | 2.11E+2 |
| HBV-M1 | 4.36E+6 | 0.248 | 0.137 | 3.2E+7 | 3.8E+2 |
| HBV-WT | 3.17E+6 | 0.228 | 0.094 | 3.36E+7 | 5.21E+4 |

**Table S2. Clinical information of 111 chronic HBV-infected patients.**

|  | **HBV (+)(n=53)** | **HBV (-)(n=58)** | ***p* Value** |
| --- | --- | --- | --- |
| Age, mean±SD years | 41.79±9.833 | 45.55±9.819 | ns |
| Gender, No. (male) | 37 | 32 | ns |
| HBV DNA, mean±SD IU/ml | 73.78±58.89 | undetected | ＜0.0001 |
| HBsAg, mean±SD IU/ml | 2011±4887 | 1598±4195 | ns |
| HBeAg, median (range) COI | 230.3(0.0933-1456) | 7.197(0.0861-201) | 0.0034 |
| ALT, median (range) U/L | 33.19(10-106) | 23.16(6-68) | 0.025 |
| AST, median (range) U/L | 31.9(14-88) | 24(9-54) | 0.004 |
| TBil, mean±SD μmol/L | 16.48±5.748 | 18.95±15.84 | ns |
